# How could a pooled testing policy have performed in managing the early stages of the COVID-19 pandemic? Results from a simulation study

**DOI:** 10.1101/2023.06.05.23290956

**Authors:** Bethany Heath, Sofía S. Villar, David S. Robertson

## Abstract

A coordinated testing policy is an essential tool for responding to emerging epidemics, as was seen with COVID-19. However, it is very difficult to agree on the best policy when there are multiple conflicting objectives. A key objective is minimising cost, which is why pooled testing (a method that involves pooling samples taken from multiple individuals and analysing this with a single diagnostic test) has been suggested. In this paper, we present results from an extensive and realistic simulation study comparing testing policies based on individually testing subjects with symptoms (a policy resembling the UK strategy at the start of the COVID-19 pandemic), individually testing subjects at random or pools of subjects randomly combined and tested. To compare these testing methods, a dynamic model compromised of a relationship network and an extended SEIR model is used. In contrast to most existing literature, testing capacity is considered as fixed and limited rather than unbounded. This paper then explores the impact of the proportion of symptomatic infections on the expected performance of testing policies. Only for less than 50% of infections being symptomatic does pooled testing outperform symptomatic testing in terms of metrics such as total infections and length of epidemic. Additionally, we present the novel feature for testing of non-compliance and perform a sensitivity analysis for different compliance assumptions. Our results suggest for the pooled testing scheme to be superior to testing symptomatic people individually, only a small proportion of the population (*>* 2%) needs to not comply with the testing procedure.

## 1 Introduction

Co-ordinated testing schemes have been a critical response tool in the COVID-19 pandemic. Testing has been key to estimating the dynamics of the new pathogen. These estimates have then been vital for nowcasting and forecasting the pandemic and so allowed for increased preparation for interventions to limit transmission [Birrell et al. [2021]]. Testing has also been necessary for its use in ‘managing’ the pandemic as identifying cases through centrally coordinated testing policies can reduce transmission and infections and minimise the cost of isolation through isolating the right people. This reduction in total and peak infections prevents illnesses especially in the context of an epidemic with only one wave (both in terms of the illness caused by the virus at the time but also the long-term effects such as long COVID), it prevents hospital admissions getting close to the hospital’s capacity and allows a faster return to normal life. In this paper, the correct identification of infected individuals given a limited testing capacity is the focus of the testing schemes. Although most of the UK’s testing scheme for COVID-19 has stopped, many countries are investing more into pandemic preparedness as scientists predict that there will be another epidemic in the near future [Gov.uk [2023b], Flanders vaccine [2023], Aboutaleb et al. [2021], Marani et al. [2021]]. Therefore, it is essential that we use what we have learnt from the COVID-19 pandemic to design an effective and efficient (in relation to a given capacity) testing scheme for these future disease outbreaks.

Due to the critical importance of testing programmes,there is a large body of literature exploring different testing schemes. The potential for the use of pooled testing (defined below) in epidemics has been suggested since its development in 1943 with increased interest during the recent COVID-19 pandemic including Statistics in Medicine hosting a discussion on whether pooled testing was ready for use in the pandemic [Dorfman [1943], Hanel and Thurner [2020], Aldridge and Ellis [2021], Citroner [2020], More et al. [2021], Haber et al. [2021a], Bilder et al. [2021], Biggerstaff [2021], Haber et al. [2021b]]. Pooled testing combines samples from multiple individuals into a single sample that can then tested together. Retesting can then be used to identify the infected individuals in the positive pools. This retesting can be done in various ways. In this paper, we use a Dorfman [1943] testing design where all individuals in positive pools are tested individually and all individuals in negative pools are marked as negative. This testing design is shown in Figure 1.

**Figure 1:**
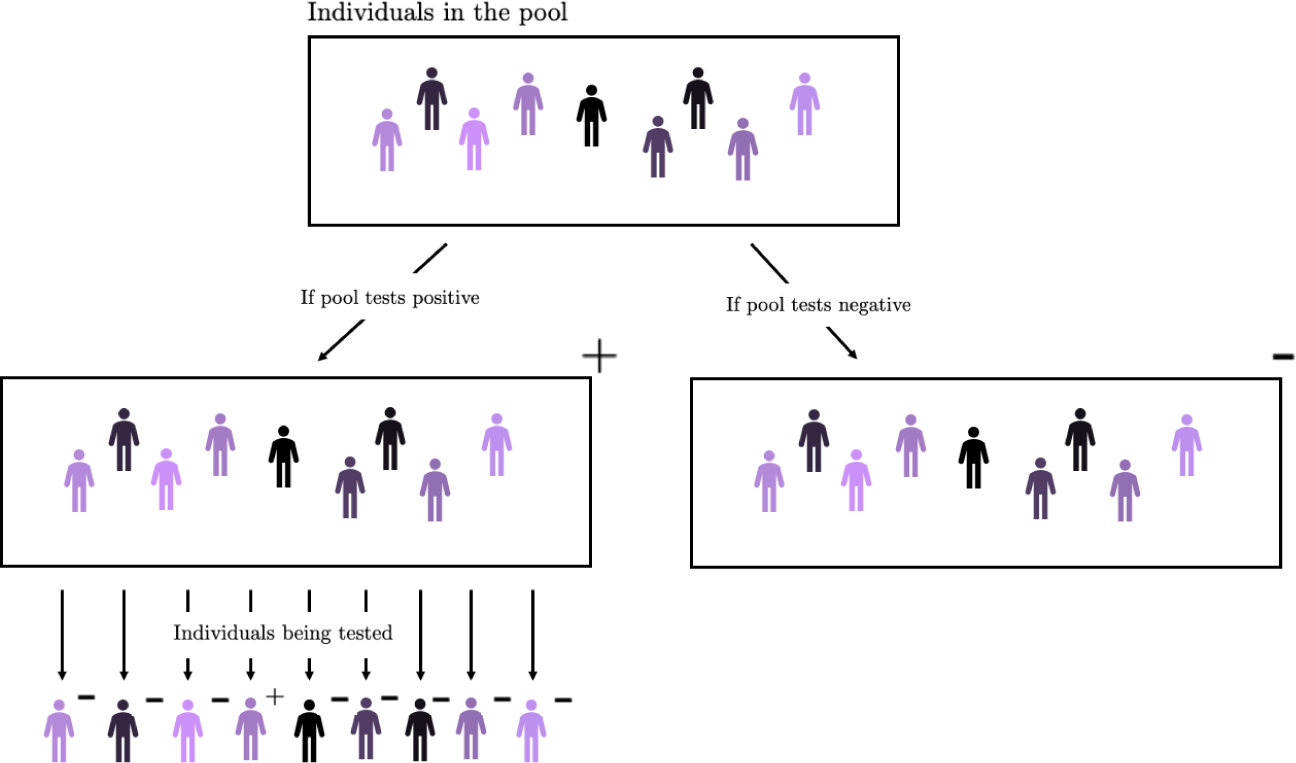
Diagram of how individuals are tested in a Dorfman [1943] pooled testing design. Tests coming back positive are marked with a ‘+’ sign and those coming back negative are marked as ‘-’. This means 10 tests (one for the pool and nine for the individuals) are used if the pooled test is positive and only one is used if the pooled test is negative

The papers suggesting the use of pooled testing for COVID-19 highlight its potential but the default comparison in the literature is individual random testing, which is randomly selecting individually for testing regardless of symptoms. However, this was not the UK’s initial testing policy in the COVID-19 pandemic, which began with only individuals who identified themselves as having symptoms being eligible for testing before the testing policy expanded to also allow targeted asymptomatic testing [Department of Health and Social Care [2020]]. Therefore, in this paper we will perform a comparison between all three of these testing methods (symptomatic individual, individual random and pooled random testing) under a number of settings.

In most of the existing literature on pooled testing (aside from a few examples such as Arasli and Ulukus [2023] who use the number of tests as a constraint), the objective of the testing scheme is to use the least number of tests possible. The issue with having the number of tests as a performance metric is that the ‘optimal’ number of tests used can exceed the number of tests per day that are available in practice. Although the number of Polymerase Chain Reaction (PCR) tests changed over time during the pandemic (varying from one PCR test for around 55 to 220 people in the UK [Gov.uk [2023a]]), the initial capacity of any future epidemic would be expected to be low and limited. Therefore, a testing scheme that requires a large number of tests is not feasible. Therefore, we want to develop a suitable testing policy to be used with a limited testing capacity as would be the case in a new epidemic.

This paper aims to develop a systematic simulation-based framework for assessing the use of different testing methods for the early stages of an epidemic. The framework starts by incorporating the dynamics of the disease of interest. In this paper we took COVID-19, a highly transmissable disease, as our motivating example. The framework then combines a detailed network that describes relationships between individuals, a realistic disease transmission model and different testing schemes that are being considered for their use in an epidemic. This framework is first used to determine which of the testing schemes is superior in terms of relevant metrics (as detailed in subsection 2.4) and then to identify properties of a disease that policy makers should be aware of when deciding on a testing scheme. For example, we consider how the proportion of infections being asymptomatic could change the preferred testing policy.

There are at least four notable contributions of our work. 1) In contrast to most existing literature, we consider the performance of testing strategies under the more realistic scenario of a limited testing capacity. This makes our results more relevant for the early stages of an epidemic than those under an unbounded capacity assumption (as a limited testing capacity would be expected at the start of an epidemic) and it allows us to suggest a wider and novel variety of performance metrics for comparing testing policies in such a context. 2) In our modelling approach, we combine a relationship network with a SEIR model that has COVID-19 specific transmission dynamics, resulting in a realistic and dynamic model to embed all testing policies considered when performing simulations. 3) We consider the relevant baseline comparator for pooled testing to be a symptomatic testing scheme rather than an individual random testing one (as commonly considered in the literature). This makes the relative comparison of pooled testing more relevant to what can currently be found in the literature. We also consider how the proportion of symptomatic infections may change the testing policy that appears superior. 4) We are, to the best of our knowledge, the first to include the impact of non-compliance into our modelling approach and to make policy makers aware which types of non-compliance are important to monitor and control. Our simulations also have the additional feature of considering errors in the testing procedure such as false negative rates.

The rest of the paper is structured as follows. In Section 2, we describe the model used to perform the simulations, detailing the relationship network, disease transmission and testing methods used. This section also contains descriptions of metrics used to analyse the different testing methods. Section 3 includes the simulation results for both the impact of the proportion of infections that are symptomatic and non-compliance. We conclude with a discussion in Section 4.

## 2 Methods

The model consists of three interconnected features: a relationship network for the model population, a disease transmission pathway (an adapted SEIR model pathway) and the testing policy. The basis of the model comes from Johnson et al. [2022], Johnson [2023], which has been adapted for realism to the COVID-19 pandemic and to study pooled testing policies. Table 1 highlights the changes from the original model, which are explored in this section. The model considers the early stages of the COVID-19 pandemic in the UK (around March 2020). There are several assumptions used in this section (such as the severity and time taken for interventions to reduce COVID transmission and uniformity in COVID transmission parameters), these assumptions can be altered by researchers in the future based on the epidemic of interest. The aim of this study is to present a systematic framework for analysing testing methods in the early stages of an epidemic.

**Table 1:** Table of comparisons between Johnson et al. [2022]’s model and our model

| Johnson et al. [2022]’s Model | Our Model |
| --- | --- |
| Social restrictions are in place throughout the model duration | Social restrictions come into effect after 14 days |
| Relationship network with transient connections and known connections (household and workplace) | Additional known connections of children going to school/nursery |
|  | Additional known social connections in the relationship network |
| Extension of SEIR where infected is split into infected asymptomatic, presymptomatic and symptomatic |  |
| All testing is done individually | Individual random, symptomatic individual and pooled random testing schemes are considered |
| Tests are completely accurate (no errors considered) | False negatives can occur |
| There is an unlimited testing capacity. | There is a limited testing capacity ($\alpha$) |
| Individuals isolate until they are no longer infected | Individuals who tests positive individually are isolated for 14 days. Individuals who are in a positive pool isolate until they can get an individual negative test up to a maximum of 14 days. |

### 2.1 Relationship Network

The relationship network represents the 1154 individuals in the model and the connections between them for a model with social restrictions and one without social restrictions. This gives a more realistic view of how COVID-19 would spread through the population rather than just having a constant probability of an individual passing on the virus to any other individual. Table 2 summarises the values used for the parameters in the relationship network. Most of these are taken to be the same as the values in Johnson et al. [2022], Johnson [2023]. One randomly-chosen realisation of the starting relationship network is used throughout the paper.

**Table 2:** Table of parameter values used in the relationship network

| Parameters in the relationship network | Their values in the relationship network |
| --- | --- |
| Number of households in the model | 500 |
| Relationship weight for known contact | 1 |
| Relationship weight for transient contact | 0.1 |
| Average number of contacts for workplace | 15 |
| Average number of contacts from school | 25 |
| Average number of social connections | 3 [Miche et al. [2013]] |
| Average number of transient connections | 10 |
| Percentage of adults working | 100 |
| Percentage of elders working | 20 |

The relationship network is set to a community of 500 households. The ‘type’ of these households (i.e., a house-hold compromised of two adults or one elderly person and three children) is randomly sampled where the probability of each household being a certain type is the proportion of households of that type in the 2011 census [Office for National Statistics [2011]]. The population in the relationship network used is 1154. Realisations of the relationship network have around 1100 individuals in them for the 500 households therefore our population size for the randomly chosen relationship network was around this average value. This population was kept quite small, due to computational cost, but was representative of the UK population.

The relationship network is set as an undirected graph with vertices representing the population and the edges representing connections between individuals. The relationships between people is split into two types of relationships: people who know each other (known contacts) and those who simply encounter each other (transient contacts). There are different ‘relationship weights’ assigned to the different types of contact with a relationship weight of 1 being assigned to known contacts and 0.1 to transient contacts. This reflects the smaller probability of transmission from transient contacts versus known contacts with the parameter values used from Johnson et al. [2022].

There are various ‘known contacts’ interactions included in the model. The most obvious are the interactions between individuals living in the same household. Individuals in the same household have edges connecting them all. Another type of known contact in the model is of connections between individuals working in the same workplace. All individuals who are in the ‘adult’ category and a fifth of those in the ‘eldery’ category have a workplace in the model. Due to this, they are connected to an average of 15 individuals via the workplace. This 15 represents the individuals in the workplace who would share facilities rather than the total size of the workplace. There are also known contacts connections between children who are in the same class. Therefore, each child has on average 24 connections with pupils in the same class as them. It is assumed that children and adults have the same progression through COVID-19, i.e., all the disease parameters that will be discussed in subsection 2.2 will be the same. This may not be a reasonable assumption for future epidemics but was the thinking at the time at the start of the COVID-19 pandemic. When using this framework for future epidemics, this assumption can be easily changed. A simplifying assumption is used that all individuals who are eligible work (i.e. all adults and one fifth of those in the ‘eldery’ category) and all children attend school/nursery and continue doing so throughout the epidemic. The model currently does not have a lockdown in place, as individuals still go to school and work, and instead have social restrictions imposed instead. Additionally, although schools were closed during the UK’s response to COVID-19, due to criticism of this policy, it cannot be assumed this would occur in a future epidemic [Leahy et al. [2021], Engzell et al. [2021], Burford [2022], Axelsson [2021]]. The optimal time for imposing lockdown, social restrictions and how these can be imposed can all be analysed using our suggested framework. Household, workplace and school connections are the known contact connections that will be in the place throughout the model.

It does not make sense for the population to stop social interactions at the beginning of the model as there are no known cases at the start. Therefore, there are two social maps in place for the model simulation. One where there are additional known social connections in the model to represent a ‘normal’ relationship network without any social restrictions and the other representing no social interactions. Each individual is connected to an average of three others by these social connections. This is based on the average number of friends that individuals in a study of 40-85 year olds had in Germany and it is assumed this would be similar to the UK [Miche et al. [2013]]. This may overestimate the connections for children in the model, as they might be expected to mainly only see those in their classes rather than additional individuals. Therefore, the model provides an upper bound on the impact of social connections. The first model with the additional social contacts is in place for the first 14 days of the model before going to the second model without any social connections. Although during the COVID-19 pandemic it took longer for the UK to introduce social restrictions, faster responses to future epidemics could be expected. A randomly selected subgraph of the population with known edge connections highlighting the additional social connections is shown in Figure 2.

**Figure 2:**
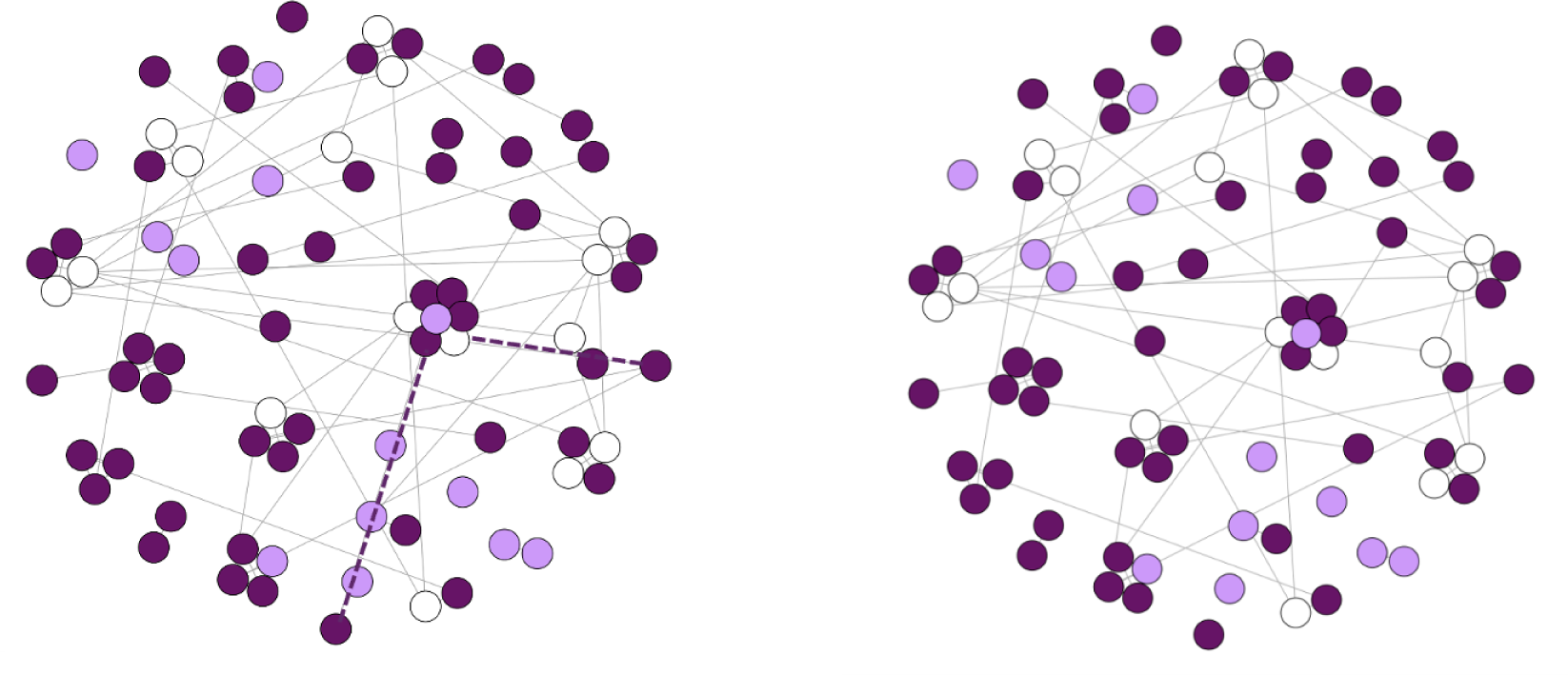
Subgraph of the population in model with edges representing known contacts. The population structure on the left is the first one in place with the additional social edges (i.e., no social restrictions); the population structure on the right is the one with fewer edges (i.e., under social restrictions). For illustration purposes, we highlight some of the edges that disappear under social restrictions. The white dots represent individuals under 18 years old, the dark purple represent individuals from 18–65 years old and the light purple dots represent individuals over 65 years old.

The transient connections represent people encountering each other doing daily activities such as travelling or shopping. There is an average of 10 transient connections per individual.

### 2.2 Disease Transmission

In this paper, we are using the example of COVID-19 to illustrate testing policies for the case of a highly transmissable emerging epidemic. Therefore, the parameter values for the transmission parameters are set for ones at the early stages of the COVID-19 pandemic [Johnson et al. [2022]]. Figure 3 illustrates the disease-state transition model used. The possible states are Susceptible, Exposed, Infected Presymptomatic, Infected Asymptomatic, Infected Symptomatic and Recovered.

**Figure 3:**
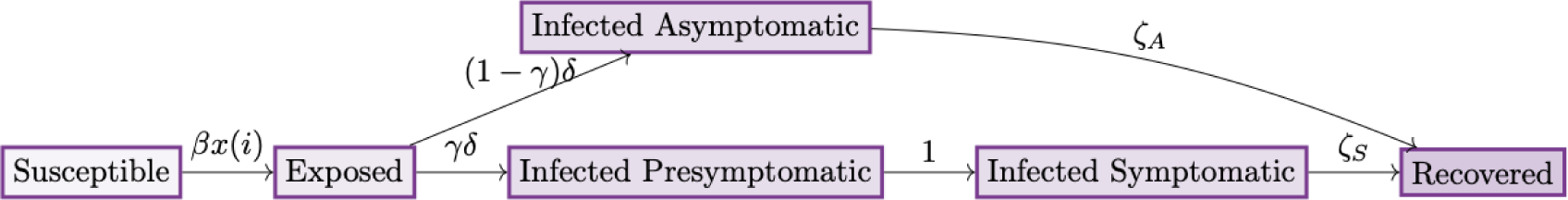
Extended SEIR model used in the paper. Arrows indicate the movement between the states with the rate of this movement written on the arrow.

Every individual starts off as susceptible apart from the initial exposed population. In our modelling, the initial outbreak consists of 20 individuals who are originally exposed cases. A sensitivity analysis was performed on the initial exposed individuals to have enough individuals exposed so the epidemic would begin but not too many that there would be a peak of infections straight away. This can be changed by other researchers using this framework and is not the main interest of this paper. Susceptible individuals have an individual risk of becoming exposed that is a function of the per-contact transmission rate, a constant denoted here as *β*, and a function dependent on the contact network for that individual *i*, denoted as *x*(*i*). Therefore, we denote the risk of susceptible individual *i* becoming exposed as *βx*(*i*). The value of *β* is set to 0.01 as used in Johnson et al. [2022].

Exposed individuals can either be asymptomatic or presymptomatic when they become infectious. An exposed individual will become infected asymptomatic with probability 1 − *γ*. This *γ* is originally set to 0.8, as was observed with COVID-19 [Buitrago-Garcia et al. [2020]]. However, this will later be varied to assess whether the conclusions still hold for varying asymptomatic proportions. The transition rate to infected asymptomatic is (1 − *γ*)*δ*, with *δ* representing the impact of the incubation period. The incubation period is modelled as a Γ(*shape* = 13.3, *rate* = 4.16) distribution [Li et al. [2020]]. The transition rate to infected pre-symptomatic is *γδ*. All infected individuals are equally likely to infect individuals they are in contact with, the only difference between infected individuals is whether they have symptoms.

Infected pre-symptomatic individuals are assumed to become infected symptomatic in exactly one day [Kucharski et al. [2020]]. In contrast, infected asymptomatic individuals remain asmyptomatic for the duration of their infectious period. The infectious period for infected asymptomatic is modelled by a (Γ(*shape* = 1.43, *rate* = 0.549) + 1) distribution [Li et al. [2020], Johnson et al. [2022]]. The duration that an individual stays in the infected symptomatic state is modelled the same as for the infected asymptomatic state minus one. These different rates for infected asymptomatic and infected symptomatic going to the recovered population are denoted as *ζ_A_* and *ζ_S_* respectively. This recovered population covers all routes out of being infectious (i.e., death and recovery) but as reinfections are not allowed in our model, these routes can be considered together.

### 2.3 Testing Schemes

This paper compares three different testing schemes: **individual random testing**, **pooled random testing** and **symptomatic individual testing** scheme.

All of the tests used in the testing schmes are assumed to be PCR tests, as these were the only tests available at the beginning of the epidemic. In a future epidemic, it is likely there will be a limited and unique form of testing. There is a lot of variation in the estimates for the false negative rate for PCR tests. We used the false negative rate for a mild case of COVID-19 as mild cases are more common than severe ones and the false negative rate is higher for mild cases than severe cases [European Centre for Disease Prevention and Control [2023]]. This puts an upper bound on the false negative rate. The false negative rate used in this paper is 27.9%, which is the false negative rate of an individual tested within the first seven days of infection for a mild case using a nasal swab [Yang et al. [2020]]. False positives were not considered, as the false positive rate for PCR testing for COVID-19 is very low [Steel and Fordham [2023]]. Therefore false positives are unlikely to have an impact on the model results. An assumption in our model is that it takes a day for a PCR test to be completed and the results returned to the individual. The different disease-testing-quarantine states that individuals in the model can be in are shown in Figure 4.

**Figure 4:**
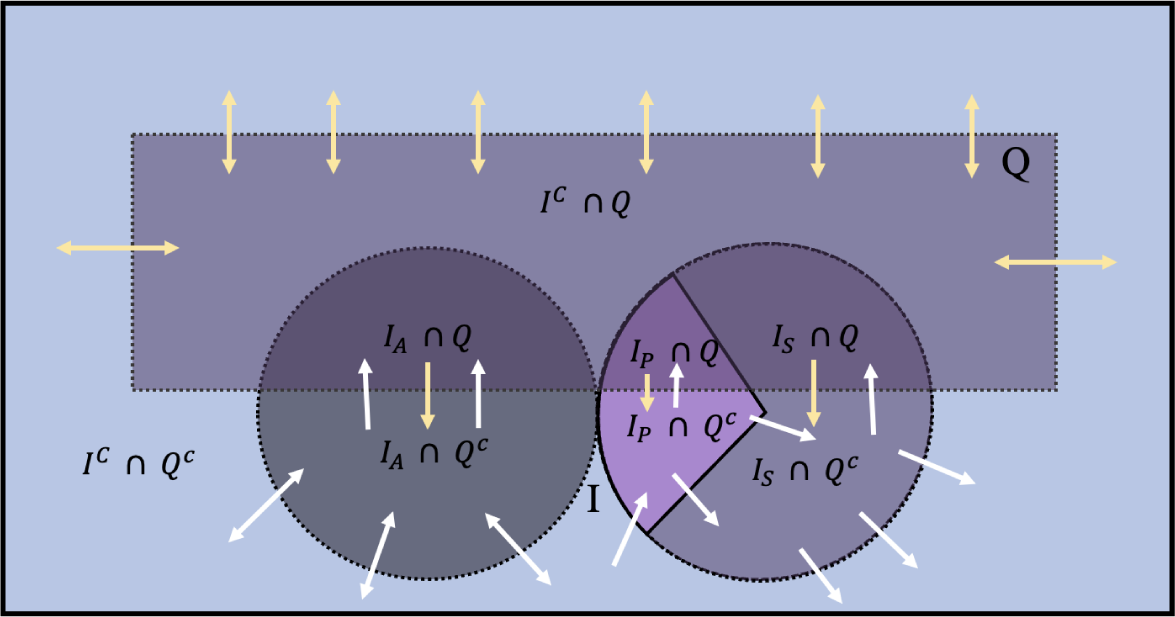
Diagram of the different disease-testing-quarantine states of individual in the model. Dotted lines represent groups that individuals can join and leave. The arrows show the directions that individuals can move through the groups. White arrows indicate movement that can happen in all testing strategies. Yellow arrows indicate directions that only happens in pooled testing.

We now introduce the notation that will be used throughout this section. Most of the notation is included in figure 4 and figure 5 but some relevant notation for testing schemes is also introduced here. Individuals in the model will move between disease-testing-quarantine states throughout the simulation. All individuals will start off as not currently isolating/quarantining and having never been isolated /quarantined (*Q^c^*) and most of the population will end up having been isolated/quarantined (*Q*) by the end of the simulation. Individuals who are currently isolating/quarantining and those who have isolated/quarantined for 14 days are considered in the same category (*Q*) as in either state an individual is ineligible for testing. Those who have completed the full 14 days are not eligible for further testing because in the individual testing setting, the individuals had to test positive to be isolated and re-infections are not permitted in the model. Individuals who have isolated for 14 days are not re-tested in the pooled testing setting to prevent individuals from being constantly put in isolation, as it would not be realistic that a person would comply with spending most of their time isolating. The infected population (*I*) is made up infected asymptomatic (*I_A_*), infected presymptomatic (*I_P_*) and infected symptomatic (*I_S_*) individuals. Depending on the testing policy in place, individuals can be put into the sample to be tested individually (*S*) or into the pooled sample (*S_P_*). If a pooled testing policy is in place, the samples from these pools (*π*) can come back positive. Individuals in positive pools (*P*) will be put into isolation/quarantine but can be removed from isolation/quarantine if their test comes back negative. Where individuals are in the model depends on both epidemic progression and on the testing scheme in place.

**Figure 5:**
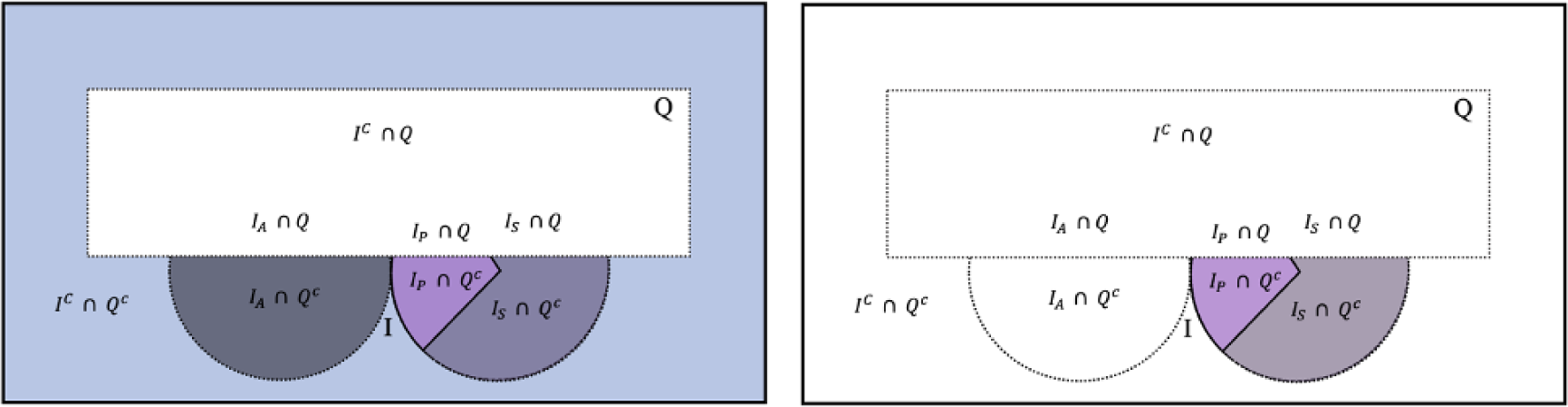
Diagram of the different disease-testing-quarantine states of the model that individuals can be sampled from. The white areas indicate areas that cannot be sampled from for testing. The figure on the left is for random testing (both pooled and individual) and the figure on the right is for symptomatic testing.

Figure 5 illustrates the main difference between the testing methods, which is the population groups (in terms of SEIR and quarantine state) that they sample from. This difference in sampling populations helps explains differences in the metrics observed in Section 3. The sampling population on the left is for random (both pooled and individual) testing and the one of the right is for symptomatic testing.

Infected and exposed individuals who are identified by these testing schemes isolate for 14 days, as was the case at the start of the pandemic [UK Health Security Agency [2020]]. Individuals who are isolating are still able to infect others in their household. However, they are unable to infect anyone outside of their household. Other infected individuals who have not been identified by the testing scheme do not change their usual activity and are able to infect their known and transient contacts. Individuals who are in a positive pool isolate until they are able to get an individual negative test result up to a maximum of 14 days. An assumption used in the model is that the total time an individual can spend in isolation in the model is 14 days. As has been discussed, this is to prevent the possibility of individuals constantly isolating. This means an individual who has already isolated for 14 days becomes ineligible for testing. This is sensible in our model, as only one wave of the epidemic is modelled, i.e., there are no re-infections, and only one pathogen is considered.

In the first part of the analysis, we assume that every individual complies perfectly with the testing scheme in place. In Section 3.2, the effect of non-compliance on these testing schemes will be considered.

#### 2.3.1 Individual Random Testing

In this testing scheme, individuals are randomly chosen from the population that are not isolating and have not previously isolated for 14 days (*Q^c^*) and then tested individually. This procedure and all following testing procedures are described using pseudocode algorithms to explain how the testing schemes were implemented. The procedure for individual random testing is described in pseudocode algorithm 1.

#### 2.3.2 Pooled Random Testing

Individuals are randomly chosen from the non-isolating and not having isolated for 14 days (*Q^c^*) population (the same population used for the individual random testing) and tested using a pooled testing scheme. The Dorfman [1943] pooled testing design is used as described in the introduction. In this testing scheme, a pre-defined number (*n_max_*) of individual samples are combined and analysed with a single diagnostic test. In what follows, we set *n_max_* = 12 as this is the maximum recommended by nhs.uk [2023] for pooling. In our model, all pools are of size *n_max_*. This should not an issue as there are not enough tests for all of the non-isolating population to be put into pools so having fewer than *n_max_* individuals and not forming another pool should not occur. For the pooled testing scheme, if needed, half of the daily number of tests are used to test individuals from positive pools. If there are not enough tests for everyone who was in a positive pool, the available tests are randomly assigned to those who were in positive pools and those not tested remain isolating up to a maximum of 14 days. The rest of the daily tests will be used to test new pools. This pooled testing design is described in the pseudocode algorithm 2. In this algorithm, the notation of ⌊*x*⌋ is used to denote the greatest integer less than or equal to *x* and ⌈*x*⌉ is used to denote the smallest integer greater than or equal to *x*. The notation of *W* (*x*) is also used where *W* (*x*) denotes the length of time individual *x* has been in isolation.

##### Algorithm 1 Individual Random Testing Algorithm

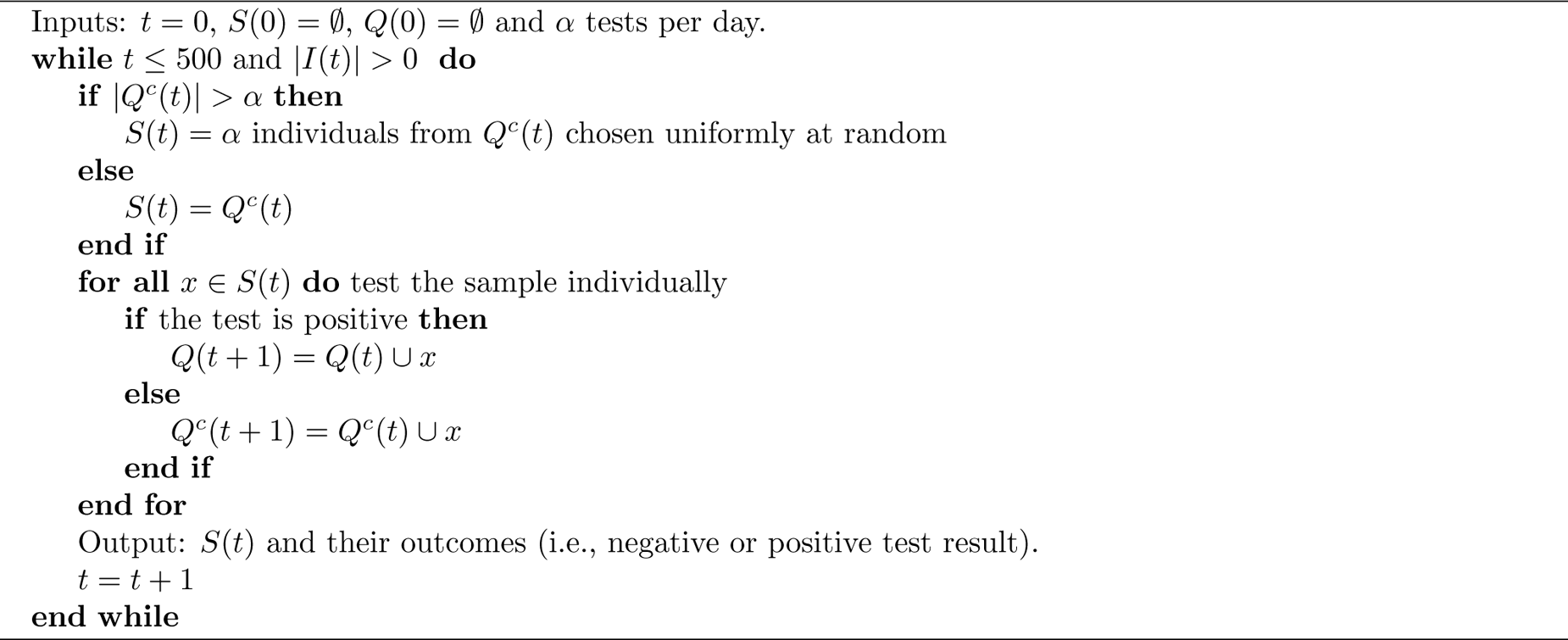

#### 2.3.3 Symptomatic Individual Testing

In the symptomatic testing scheme, only individuals who are in the (infected) symptomatic state (*I_S_*∩ *Q^c^*) are eligible for testing. In our model, as it is assumed there are no other infections with similar symptoms to COVID-19, initially all individuals who are symptomatic are infected. This assumption will be relaxed in subSection 3.2. They are then tested individually for as many as the capacity allows for. Remaining unused testing capacity is used to perform individual random testing on the non-isolated individuals (*Q^c^*). The symptomatic testing is described in the pseudocode algorithm 3.

### 2.4 Metrics to be considered

As discussed previously, unlike the vast majority of papers in the literature, we will not consider the performance metric of how many tests are used in the testing procedure. There is a small and fixed testing capacity therefore it would be expected that the number of tests used would always be close to capacity (except from at the start and end of the pandemic) and thus would be not be informative of scheme differences. As done by Arasli and Ulukus [2023], we instead fix the number of tests per day and measure other key performance metrics. The number of tests per day (*α*) took the values of 5, 10, 15 and 20. This is because when we compare the UK’s testing capacity during the pandemic and scale it down proportionally to our the population of 1154, we find the number of tests per day never goes above 20 [Gov.uk [2023a]].

The other metrics that we report are:

- The expected total number of infections. This is the total number of individuals who are infected over the length of the epidemic (i.e., before no more infections occur).
- The expected total isolation period. This is the total number of days spent in isolation for the whole of the population,

#### Algorithm 2 Pooled Random Testing Algorithm

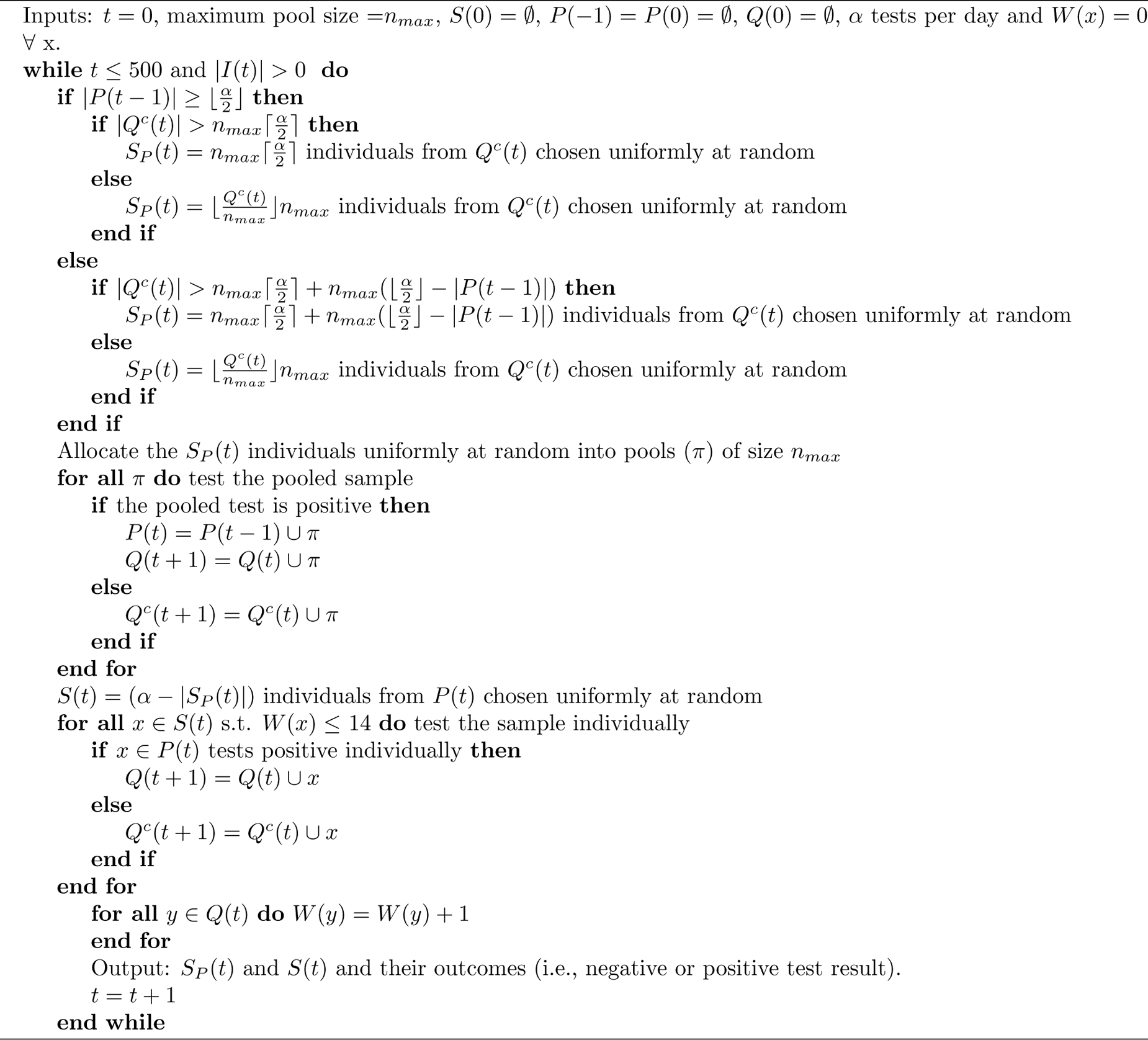

#### Algorithm 3 Symptomatic Individual Testing Algorithm

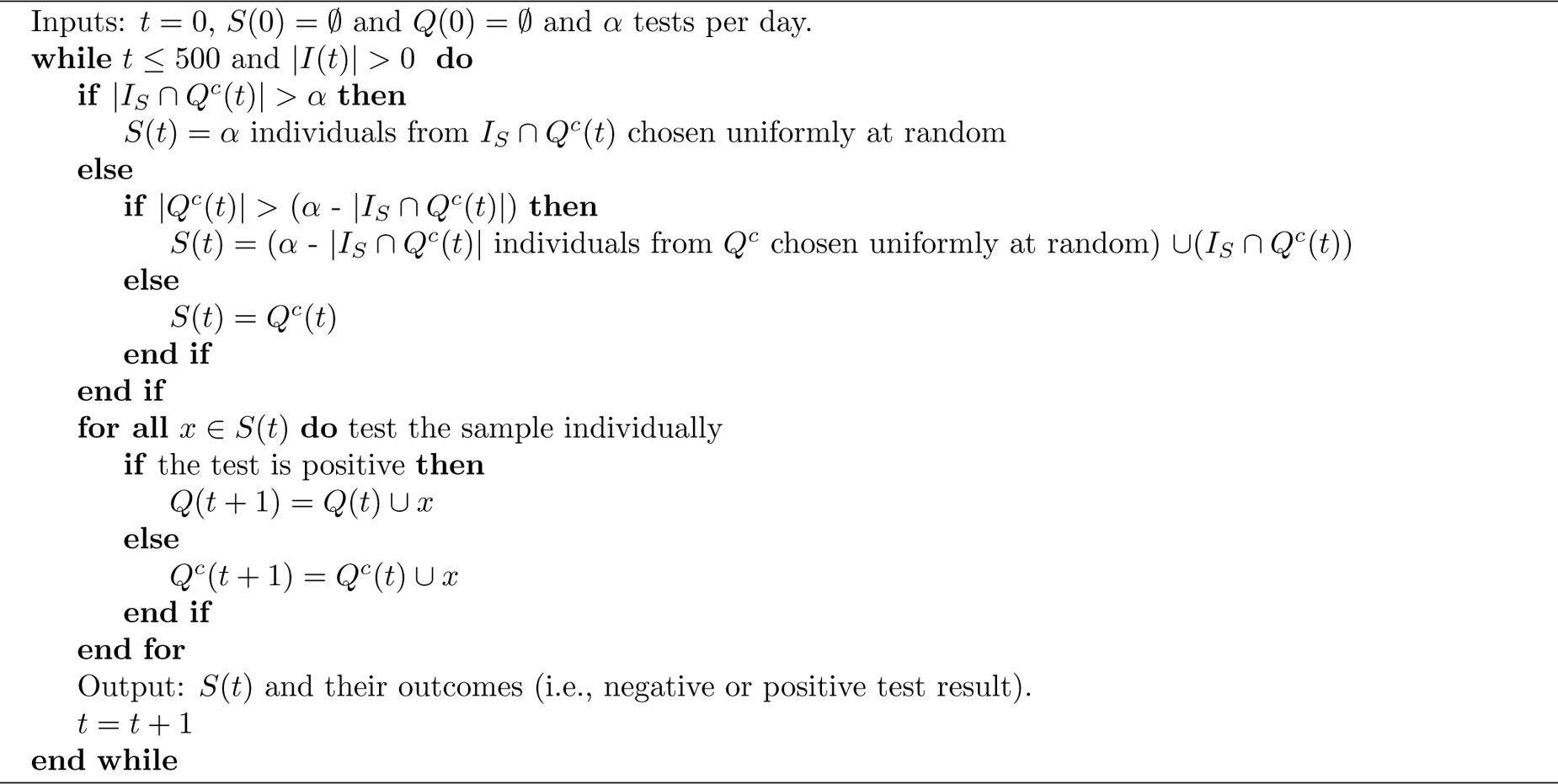

- The expected length of the epidemic. The total time from beginning of the simulation until no new infections occur,
- The expected peak size. The greatest number of individuals infected at once,
- The expected total number of days that the new infections crosses a certain threshold of infections per day. This threshold was set to 5 and is the equivalent of when the UK first when into lockdown based on the population of 1154 compared to the UK’s entire population [Birrell et al. [2021]].

## 3 Simulation

The simulation operates on a time unit of a day and lasts until there are no more infected individuals (with a cap of 500 days). The epidemic is always controlled in the model as there is a fixed number of individuals in the population (i.e., no-one is entering or leaving) and there are no reinfections or other infections with similar symptoms. This means in every simulation run eventually enough of the individuals in the model have been infected so that the probability that an infected individual is in contact with a susceptible individual is so small that spread stops. This represents the first wave of an epidemic. Even though there were multiple waves in the case of COVID-19, this may not be the case for future pandemics.

All simulations are run based on one contact network to allow ease of comparison. To get each expected value of a metric, the model is run 200 times and the mean values across these model runs are reported. The simulation begins with 20 people being exposed to COVID-19 (just less than 2% of the population).

### 3.1 Varying Symptomatic Proportion

Initially, the model has 80% of infected people having symptoms, as was the case for the original strain of COVID-19 [Buitrago-Garcia et al. [2020]]. However, this was not the case for other COVID-19 variants and may not be the case for future pandemics especially if the symptoms are less noticeable or verifiable. There is also variation in the estimated proportion of symptomatic infections. For example, the 95% confidence interval for Buitrago-Garcia et al. [2020]’s estimate for the first variant is [75,83]. We varied the proportion of symptomatics from 80% to perform a sensitivity analysis of this parameter on the relative comparisons of the testing strategies. The symptomatic proportion is set as low as 0% for illustrative purposes, which for the symptomatic testing scheme means there is no testing. Therefore, we can see how each policy differs from no testing. The performance metric of expected total number of infections is shown in Figure 6.

**Figure 6:**
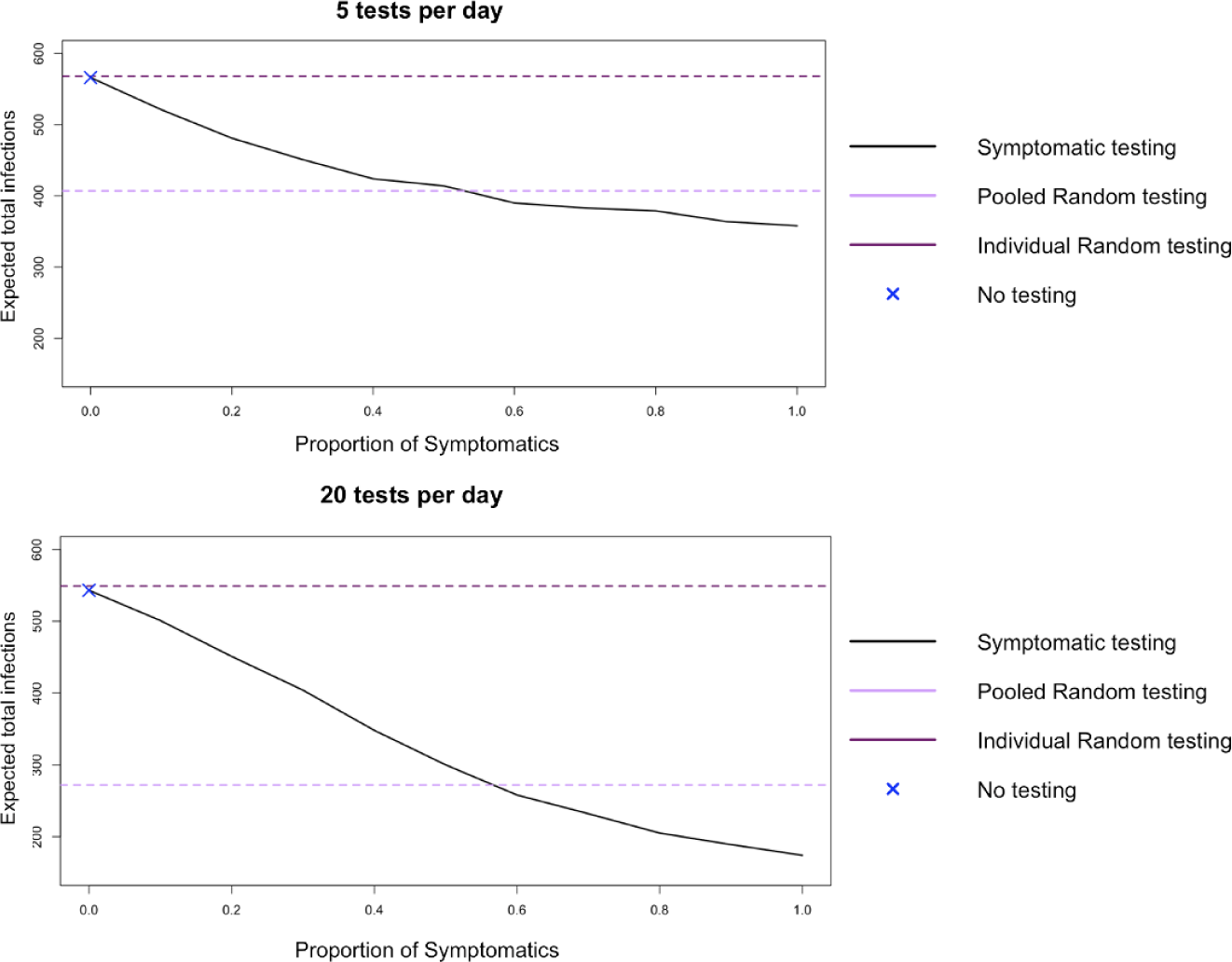
Graphs of proportion of symptomatics against the expected total number of infections. The top graph is for 5 tests per day and the bottom one is for 20 tests per day. No testing corresponds to the symptomatic testing policy with the proportion of symptomatics being 0%.

We see that in both of these settings (5 tests per day and 20 tests per day) for a high-medium proportion of symptomatics (i.e., over 50% symptomatics), the expected total infections is less for symptomatic testing compared with pooled testing. This provides evidence that for the first variant of COVID-19, the implemented policy of symptomatic testing was a good one in terms of reducing the total number of infections. However when the proportion of symptomatics is lower (less than 50%), pooled testing becomes the best policy. We note that pooled individual testing clearly outperforms individual random testing and therefore we can dismiss this as an alternate testing policy for this setting.

We also see that the expected total infections for individual random testing is only slightly less than no testing. This is because infections are very unlikely to be identified particularly in this scenario of a dynamic model where the individuals who are infected are constantly changing.

The next performance metric to be considered is the expected length of the epidemic. The length of the epidemic is the amount of time from when the first individual in the model is infected until the last individual in the model in infected. The graph of length of epidemic for varying proportion of symptomatics is figure 7.

**Figure 7:**
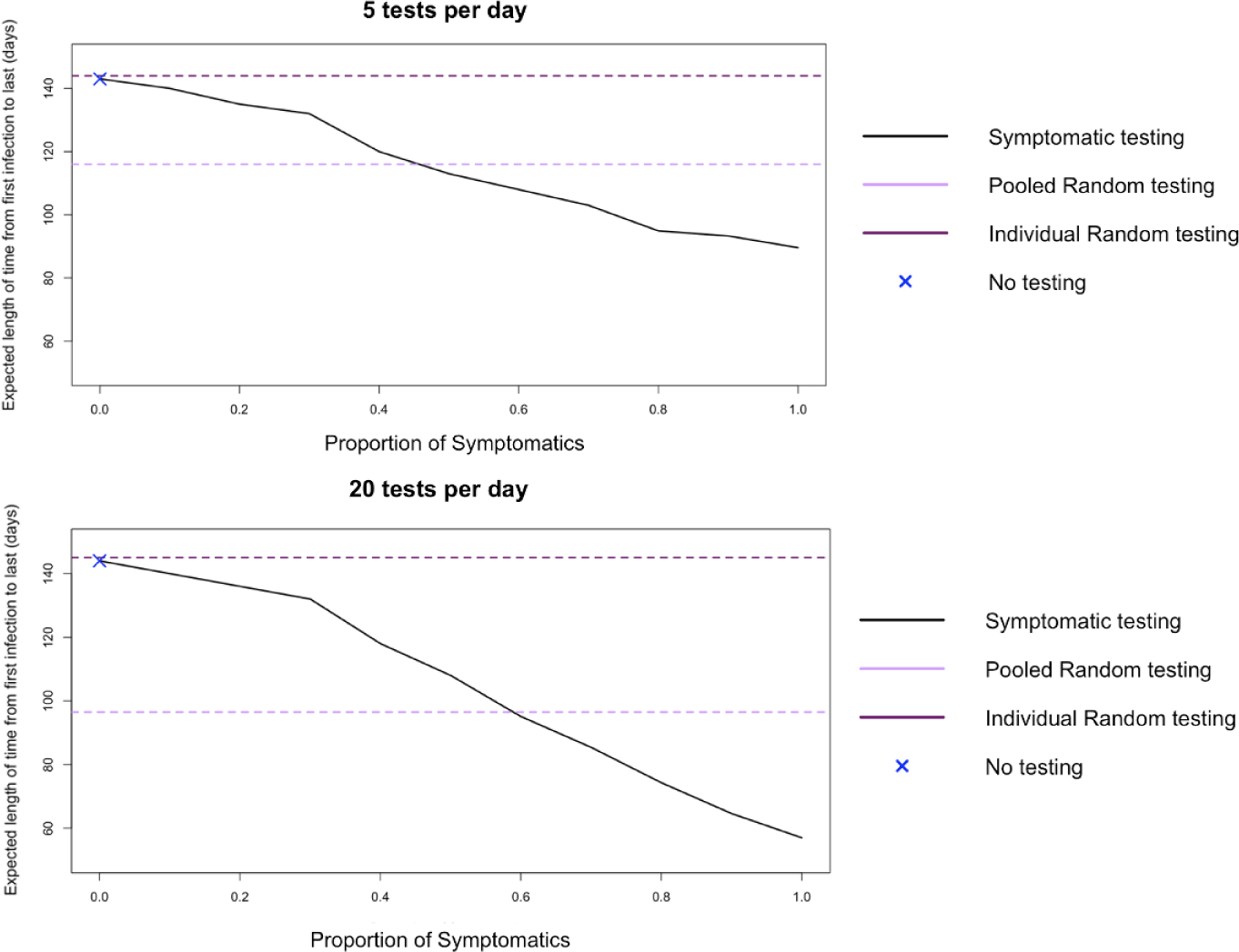
Graphs of proportion of symptomatics against the expected length of the outbreak. The top graph is for 5 tests per day and the bottom one is for 20 tests per day. No testing corresponds to the symptomatic testing policy with the proportion of symptomatics being 0%.

As we have seen for the performance metric of expected total number of infections, there is a similar pattern where pooled testing performs better up to about 50% of cases being symptomatic and for symptomatic rates greater than 50% symptomatic testing performs better. We also see the crossover point at which pooled testing performs better is higher for a greater number of tests per day.

When we consider pooled testing we see two main features. The first is it is not good at identifying cases quickly enough to prevent a large peak. As shown in Figure 8, the cross-over point for when symptomatic testing performs better at reducing the size of the peak is only around 30% of cases being symptomatic. This could be because the testing policy selects randomly from the non-isolating population so the probability that an infected person is selected for the pool in the initial stages of their infection is low.

**Figure 8:**
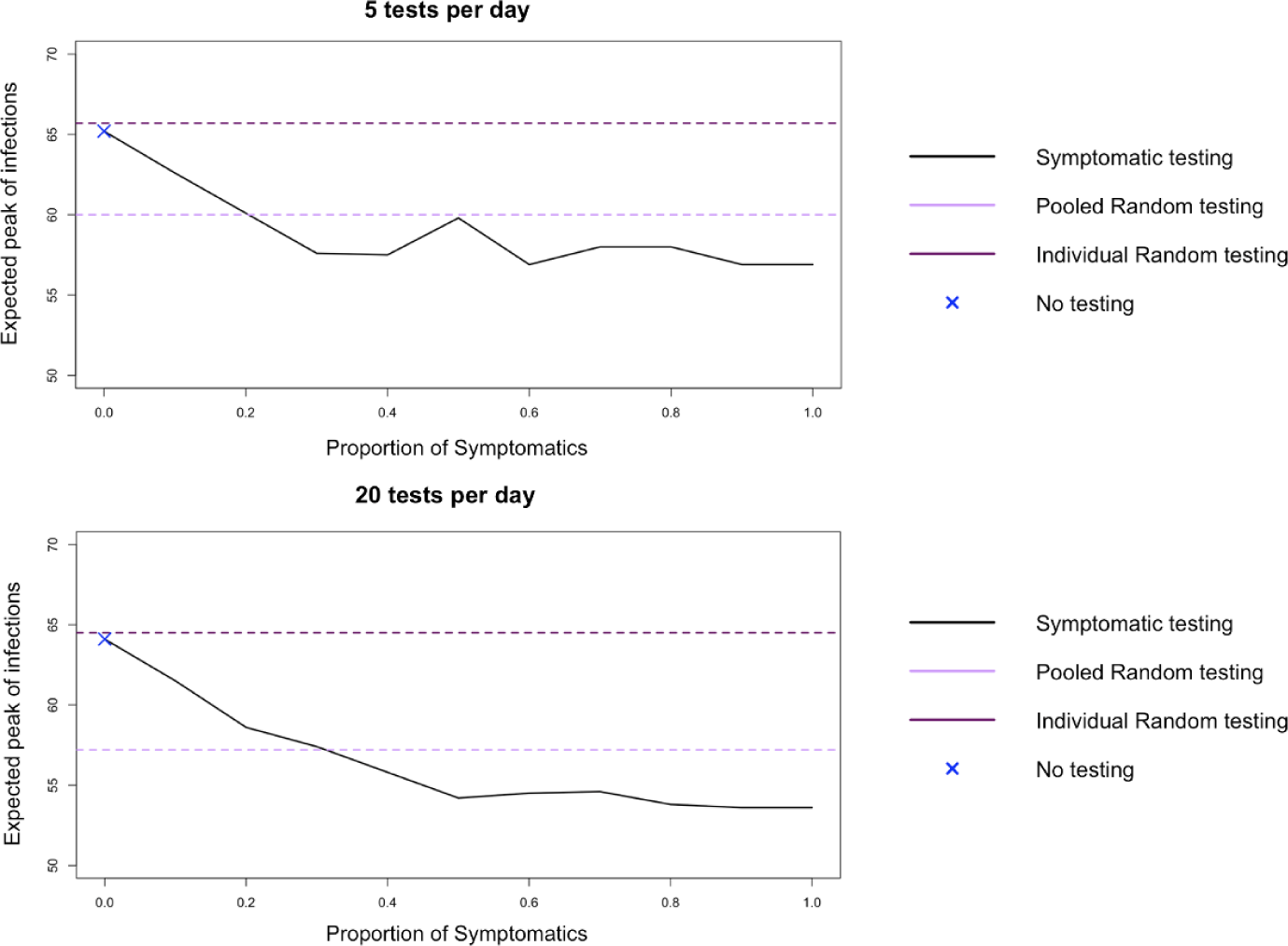
Graphs of proportion of symptomatics against the expected peak size for each of the three testing methods. The top graph is for 5 tests per day and the bottom one is for 20 tests per day. No testing corresponds to the symptomatic testing policy with the proportion of symptomatics being 0%.

We can see that individual random testing performs similarly to no testing when it comes to reducing the expected size of a peak, as the individual random testing is very unlikely to identify infected individuals. The slight difference in the expected peak size between individual random and no testing is due to statistical variation. Although these increase in peak sizes may be small, it is only for a population size of 1154. When we consider that a catchment size of a hospital could be about 618 times this, the difference in peak size becomes substantial [Beaney et al. [2022]].

The second notable feature of pooled testing is the increase in the total isolations. As individuals who are in positive pools must isolate until they have a negative individual test, which takes a day to be done even when there is capacity for this testing, this means individuals have to isolate unnecessarily. Indeed uneccesary isolations can account for about 75% of the isolations that occur in the pooled testing design. For the case of five tests per day and for medium to high proportion of symptomatics, there is an average of about two days in isolation per individual in the model for symptomatic testing but for pooled testing this increases to about 5 days. This pattern holds for larger number of tests per day, with the average number of days in isolation for the pooled testing scheme being about three times that of the symptomatic testing one with this difference increasing as the number of tests per day increases. The difference in total isolations for the case of 5 tests per day is contained in table 3 in the appendix.

**Table 3:**
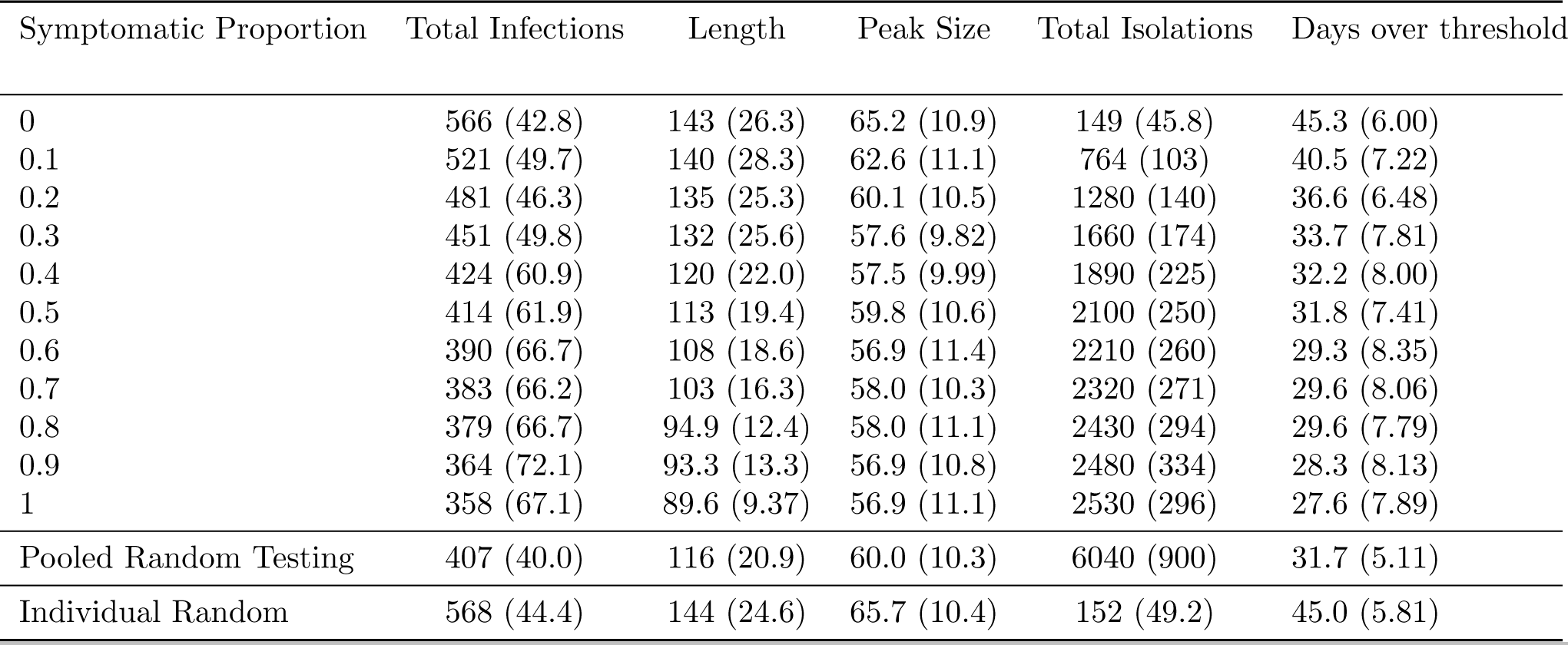
Table of the expected value and standard deviation of total infections, length of epidemic, peak size, total isolations and the number of days the number of new infections surpasses five for symptomatic testing for varying symptomatic proportions and with pooled testing and individual random testing as comparison for 200 replicates and for 5 tests per day

| Symptomatic Proportion | Total Infections | Length | Peak Size | Total Isolations | Days over threshold |
| --- | --- | --- | --- | --- | --- |
| 0 | 566 (42.8) | 143 (26.3) | 65.2 (10.9) | 149 (45.8) | 45.3 (6.00) |
| 0.1 | 521 (49.7) | 140 (28.3) | 62.6 (11.1) | 764 (103) | 40.5 (7.22) |
| 0.2 | 481 (46.3) | 135 (25.3) | 60.1 (10.5) | 1280 (140) | 36.6 (6.48) |
| 0.3 | 451 (49.8) | 132 (25.6) | 57.6 (9.82) | 1660 (174) | 33.7 (7.81) |
| 0.4 | 424 (60.9) | 120 (22.0) | 57.5 (9.99) | 1890 (225) | 32.2 (8.00) |
| 0.5 | 414 (61.9) | 113 (19.4) | 59.8 (10.6) | 2100 (250) | 31.8 (7.41) |
| 0.6 | 390 (66.7) | 108 (18.6) | 56.9 (11.4) | 2210 (260) | 29.3 (8.35) |
| 0.7 | 383 (66.2) | 103 (16.3) | 58.0 (10.3) | 2320 (271) | 29.6 (8.06) |
| 0.8 | 379 (66.7) | 94.9 (12.4) | 58.0 (11.1) | 2430 (294) | 29.6 (7.79) |
| 0.9 | 364 (72.1) | 93.3 (13.3) | 56.9 (10.8) | 2480 (334) | 28.3 (8.13) |
| 1 | 358 (67.1) | 89.6 (9.37) | 56.9 (11.1) | 2530 (296) | 27.6 (7.89) |
| Pooled Random Testing | 407 (40.0) | 116 (20.9) | 60.0 (10.3) | 6040 (900) | 31.7 (5.11) |
| Individual Random | 568 (44.4) | 144 (24.6) | 65.7 (10.4) | 152 (49.2) | 45.0 (5.81) |

We can see the difference that the proportion of symptomatic makes when we consider multiple performance metrics together: the expected length of epidemic, total expected number of infections, expected number of days that the number of new infections exceeds five, total expected isolation days and expected peak size. Radar plots of these five metrics for the different testing schemes are shown in figure 9.

**Figure 9:**
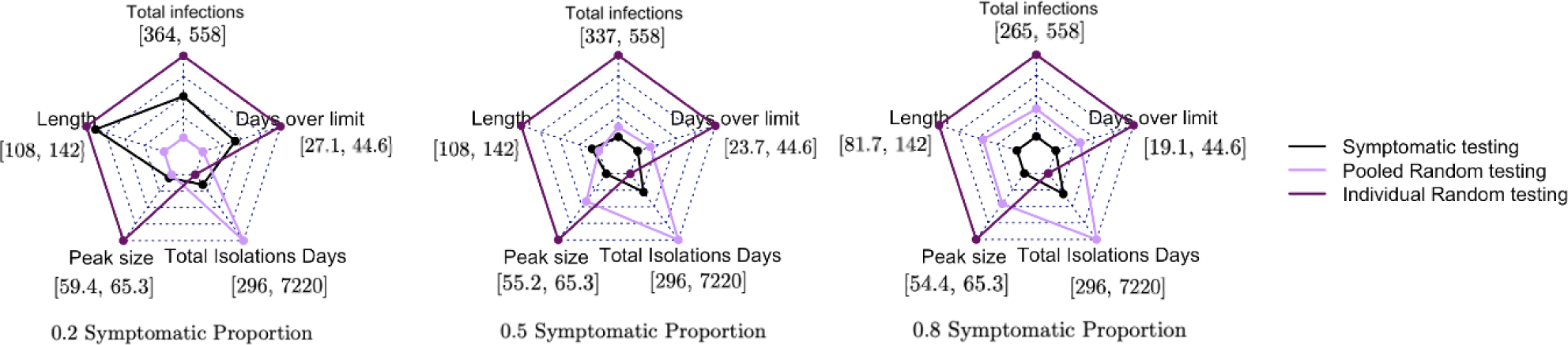
Radar plots for the expected values of five performance metrics for different symptomatic proportions (0.2 on the left, 0.5 in the middle and 0.8 on the right) for the three testing schemes for 10 tests per day. The range for each performance metric is written in square brackets. For reference, there was 1154 people in the model and the range of length of infections across all simulations was [57.0, 147]. The closer the point is to the centre, the better that testing policy does for that metric.

In each diagram the pooled random testing and individual random testing are the same for each symptomatic proportion, as these testing schemes do not depend on the symptomatic proportion. The difference in them that can been seen in figure 9 is due to the changing comparison to the symptomatic individual testing. We see for a low proportion of symptomatic infections that pooled testing appears better. For a medium or high symptomatic proportion of symptomatic infections, symptomatic testing appears better for most of the metrics considered. As only for a low proportion of symptomatics does the proportion of symptomatics affect the better testing policy, in the rest of the analysis we will use a symptomatic proportion of 0.8.

### 3.2 Non-Compliance

Non-compliance is an important feature in the model, since (as will be shown) it can have a dramatic impact on which is the best testing policy to use. It also adds realism to the model, as a testing policy where every individual in the model perfectly complies with all testing policies is unrealistic. Despite this, to the best of knowledge, we are the first to consider non-compliance. In what follows, we again use the parameters for COVID-19, including a symptomatic proportion of 0.8.

Three types of non-compliance and also the difficulty of distinguishing symptoms are considered in the model. These are:

1. Individuals refusing to participate in any testing schemes or isolate regardless of whether or not they are infected.
2. Individuals who are symptomatic (who should therefore go to get a test done in the symptomatic testing case) instead wait one day before going to get tested. This could be for a range of reasons such as to see if their symptoms improve, they do not have time to get a test or they do not think their symptoms are severe enough.
3. Individuals who do not have symptoms but say that they have symptoms to be eligible for testing under a symptomatic testing scheme. Individuals who have symptoms of COVID-19 but not COVID-19 (i.e., they could have another disease) impact the system in the same way as this non-compliance, as testing capacity is used for individuals not infected with COVID-19. Therefore, this type of non-compliance and the issue of distinguishing disease symptoms will be considered together.

The first of these considered is the proportion of individuals who refuse to participate in any testing or isolation scheme. The radar plots for the effect of this non-compliance are contained in figure 12 in the appendix. We see that symptomatic testing outperforms pooled testing across all performance metrics as was the case when there was no non-compliers. This is because there is the same number of individuals not complying with all of the methods so should affect them all similarly.

We will now consider the other two types of non-compliance. Both of these only affect the symptomatic testing scheme, as this is the only one that requires individuals to put themselves forward for testing rather than being randomly selected from the population. The first type concerning symptomatic individuals waiting before getting a test is modelled by varying the probability that a symptomatic person will wait a day before getting tested. There is no limit on the number of days that a symptomatic person can choose to wait a day (although eventually the individual will recover and no longer need to be tested). The other type of non-compliance is modelled by varying the proportion of the population that put themselves forward to have symptomatic testing even though they are not in the infected symptomatic state. This is capped at 10% both due to pooled testing already looking so much better but also as it would be unlikely that over 10% of the population would falsely claim to have symptoms and the social restrictions in the model should reduce the spread of other diseases with similar symptoms. The effect of these two types of non-compliance on expected total number of infections is shown in the heat map in Figure 10.

**Figure 10:**
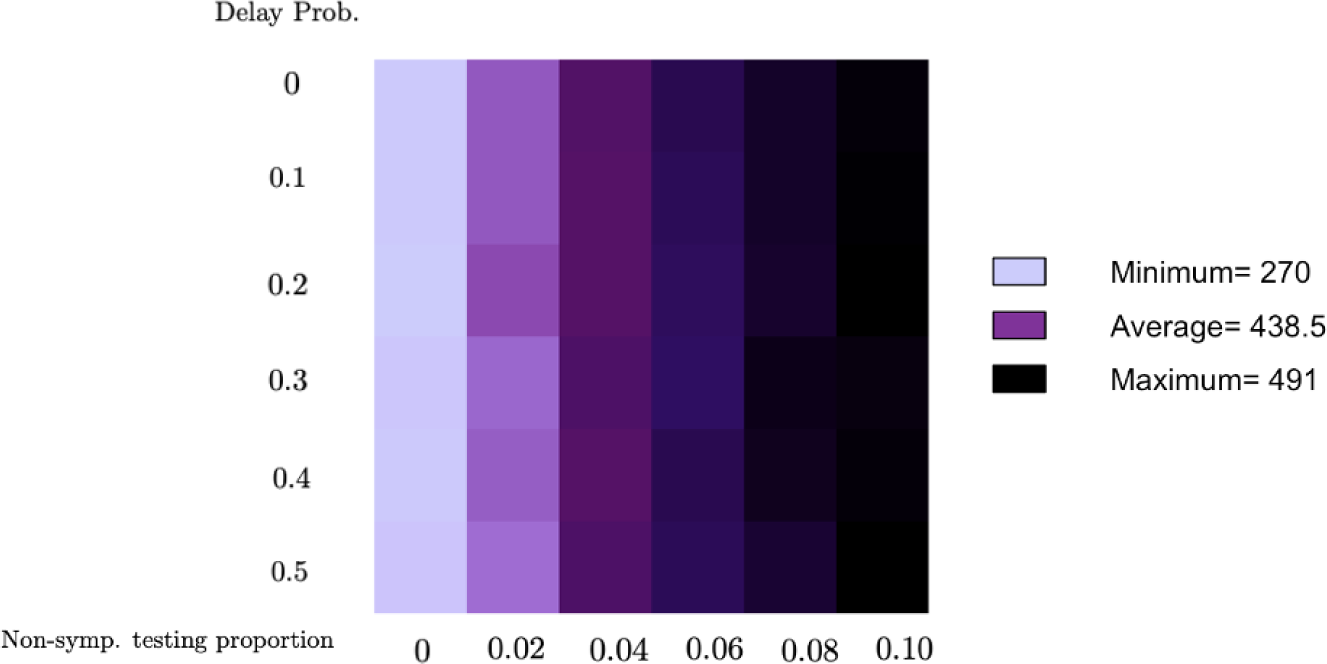
Heat map of expected total number of infections for symptomatic testing when varying both the probability of individuals delaying a day before getting tested and the proportion of the population that do not have symptoms but still go to get tested under the symptomatic testing scheme for 10 tests per day. For comparison, the total number of infections for pooled random testing is 360.

Although the probability of a symptomatic individual delaying testing by a day slightly increases the total expected number of infected individuals, we see that this does not cause a large increase in the expected total number of infections. This is because most individuals have to wait a day anyway as there are not enough tests. Therefore whether the individual chooses to delay a day or the lack of testing capacity means an individual has to delay a day, it makes little effect on the total expected number of infections. However, the other parameter of the proportion of the non-symptomatic population that go to get tested clearly makes a large difference. This is because even if only 2% choose to get tested then this is about 20 extra individuals who are competing with the symptomatic population for 10 tests per day. Therefore, as this proportion increases, the probability that the tests are given to the ‘correct’ individuals decreases.

The decision tree in Figure 11 demonstrates the conditions for each testing policy to perform better in terms of the metric of total infections. The decision tree for length of epidemic would be the same.

**Figure 11:**
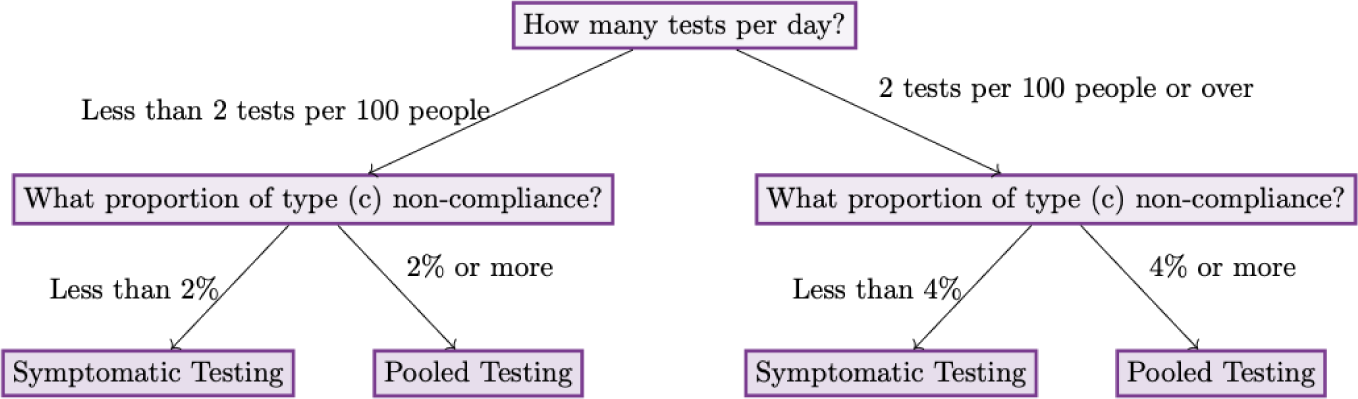
Decision tree for testing policy under certain non-compliance conditions for non-compliance of type (c) as defined earlier in this section for the metric of total infections

**Figure 12:**
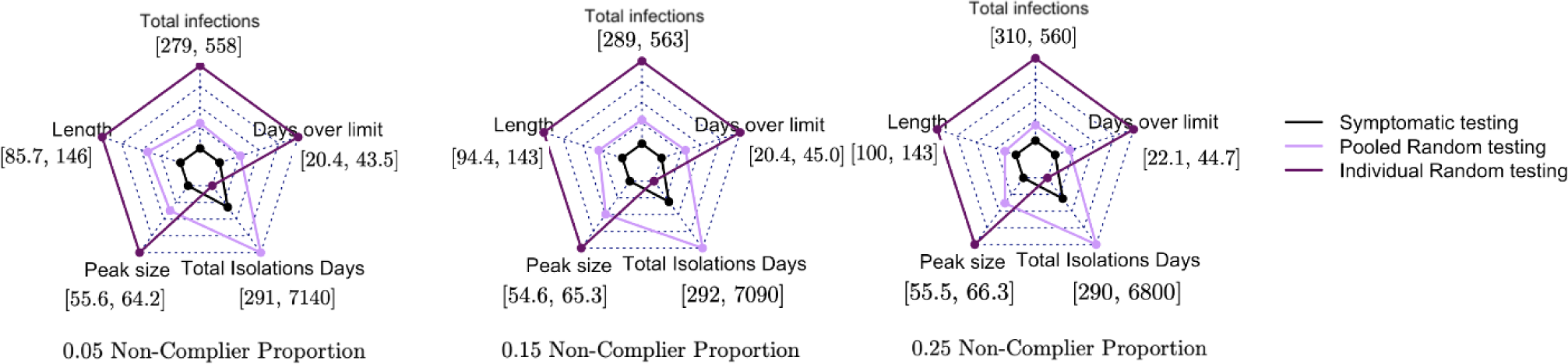
Radar plots for the expected value of five performance metrics for different proportions of individuals not complying with any testing scheme - 0.05 on the left, 0.15 in the middle and 0.25 on the right - for the three testing schemes for 10 tests per day. The range for each performance metric is written in square brackets. For reference, there was 1100 people in the model and the range of length of infections across all simulations was [57.0, 147]. The closer the point is to the centre, the better that testing policy does for that metric.

This demonstrates that the proportion of individuals who do not have symptoms but still get tested is very important when deciding which of the testing schemes to use. An alternative way of thinking of about this is if we want our symptomatic testing scheme to be successful, it is critically important to have a policy that limits individuals who are not symptomatic being able to access tests that are only meant for symptomatic individuals. Indeed it is more important to understand the level of non-compliance than it is to understand the proportion of symptomatics when it comes to choosing a policy, as non-compliance has a much greater effect on choosing the best policy. This was not a large issue for COVID-19 as most symptoms (such as fever) were clearly observable; however this may not be the case for a future epidemic. Figures 13 and 14, contained in the appendix, are the decision trees for days over threshold of 5 new infections and peak size. If the metric of interest is days in isolation then symptomatic testing should be chosen in all settings.

**Figure 13:**
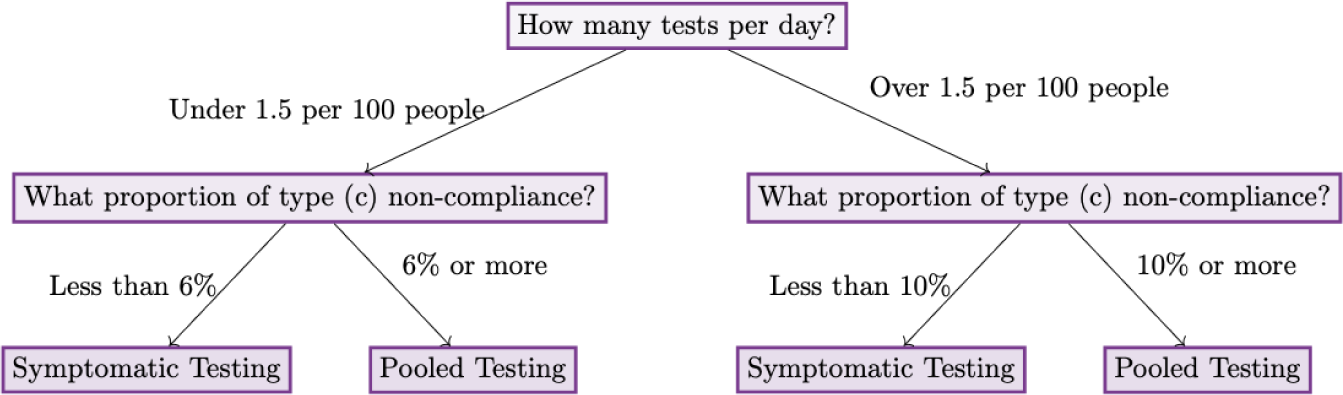
Decision tree for testing policy under certain non-compliance conditions for non-compliance of type (c) as defined in *section 3.2* for the metric of peak size

**Figure 14:**
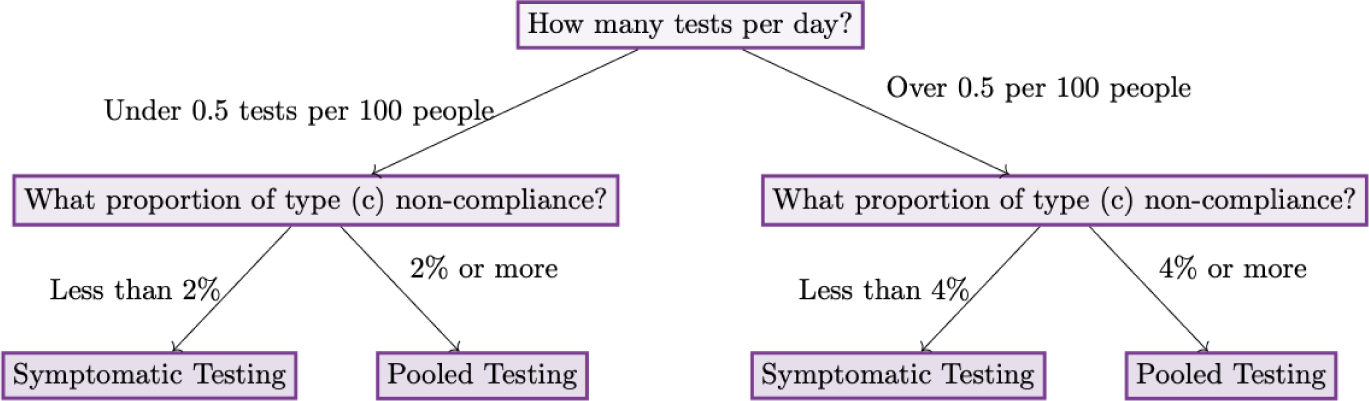
Decision tree for testing policy under certain non-compliance conditions for non-compliance of type (c) as defined in *section 3.2* for the metric of days new infections surpass the threshold of five

## 4 Discussion

In this paper, we developed a systematic framework to analyse the performance of different testing methods in the context of the early stages of an epidemic using the dynamics of COVID-19 as an example. Our framework consists of a realistic model for the pathogen, inclusion of testing features that represent PCR tests as accurately as possible, features of the population such as different types of non-compliance and a broad enumeration of critical metrics.

In contrast to most existing literature, our model incorporates a fixed number of tests per day and reports other performance metrics. This means a more accurate modelling approach of the best testing policy, as the number of tests used cannot exceed the capacity that was available during the pandemic. This means the reductions in other performance metrics (such as the total number of infections and peak size) are actually achievable under limited testing as would be the case at the beginning of an epidemic.

We showed through the example of the dynamics of COVID-19 that symptomatic testing is an effective way of controlling an epidemic in the context of the early stages of an epidemic when a medium or high proportion of cases are symptomatic and there is a good level of compliance. If less infections are symptomatic then it is clear that a pooled testing policy should be used instead. We also used individual random testing as a comparison as has been done in many papers in the literature. We saw that pooled random testing does outperform individual random testing. However, when also considering symptomatic testing we see that individual random testing is not the best baseline comparator to use for testing schemes. Although individual random testing does not perform very well in managing an epidemic, it can still be very useful for surveillance.

This paper also illustrates the importance of considering non-compliance in models when assessing the value of testing schemes as a public health policy. This not only adds realism but also indicates which types of non-compliance policymakers should be concerned about when selecting a policy (and to which extent). In our COVID-19 example, we saw if a lot of individuals who did not truly have symptoms went for testing, then symptomatic testing no longer appears to be the best policy to be used. This indicates to policy makers the necessity of having some sort of check when implementing symptomatic testing to make sure that those that turn up for the testing truly have symptoms. It also highlights that a symptomatic testing policy does not make sense when dealing with an illness that does not have easily verifiable or unique symptoms, such as the only symptom being tiredness, as it would be impossible to ascertain which individual’s assessment of their own symptoms are in fact accurate. Although these individuals (and individuals infected by other pathogens) are being compliant, as they are coming in for testing when they have symptoms, this is not desirable. Under social restrictions, this should be less of an issue as the spread of other diseases would be hindered however common symptoms, like tiredness or headaches, would still be present. However if we want to consider testing policies as we transition back to normal life, the prevalence of other diseases with similar symptoms would also have to be seriously considered.

The model is currently designed for its use in diseases that spread through respiratory droplets. Although, this model can be used for other diseases that do not spread this way (e.g., HIV), the relationship network would need to be amended to illustrate this type of transmission as well as the parameters for disease transmission. Equally, the paper considers the case of the early stages of an epidemic. However, this can be expanded using a similar framework to the one developed to consider what is the best testing policy for different phases of an epidemic such as the transition back to ‘normal life’. Different metrics would need to be considered but a similar analysis can be produced. In the transition back to ‘normal life’, it would also be unlikely that there would be social restrictions and hence the model could be adapted to keep the first version of the relationship network throughout the simulation run. The other consideration for later on in an epidemic is whether other quicker/cheaper forms of testing have been developed that can be used in the pooled testing procedure. In work in progress, we are considering the use of Lateral Flow Devices (LFDs) for the follow-up testing for individuals in positive pools and the benefits and drawbacks of this in the context of a UK hospital setting.

There are many assumptions made in the modelling (such as the type and speed of disease transmission mitigation measures and the uniformity of disease transmission parameters used) however these can easily be amended and, indeed, the model framework can be used to assess these interventions. For a future epidemic, the framework would need to be adapted to add the estimates of pathogen parameters, testing policies that are being considered, estimates of compliance and the interventions measures in place but then it can be used to assess testing policy or any other policy.

One limitation of the current study is that it only considers a population of 1154. The concern would be that the conclusions drawn based on that population would not hold for other population sizes, particularly when suggesting policy for the whole of the UK. Although it would be best to run a larger scale simulation, these larger scale simulations quickly become infeasible due to their computational burden. However, we note that most of the UK is made up of communities like the ones considered in our model. Therefore, if the conclusions are relevant for the scale of the community and the UK is made up of communities like these then it should be relevant for the UK.

Another limitation of our study is that pooled testing is being used consistently throughout the course of each simulation run. This is terms of the pool size, where we have used the NHS’ recommended pool size of 12. This pool size is also consistent across all groups in the model. Filiatreau et al. [2022] have found efficiency gains in pooled testing by having different pool sizes for different groups depending on the group’s probability of testing positive. Therefore, it would be beneficial to see if these efficiency gains remain in the case of a limited test capacity. The pooled testing design was also consistent in terms of at least half of the pools being used for testing new pools and half being used for re-testing individuals. This is not optimised in accordance to any criteria. Therefore, there could be gains made in certain performance metrics if another split of testing new pools versus retesting individuals in positive pools is used. Alternatively, this split could be varied throughout the model depending on prevalence. However, we must consider when we are optimising that there is a trade-off between limiting time in isolation and limiting transmission of the virus, an issue that has been highlighted by researchers [Fearon et al. [2021]].

The model currently does not consider metrics such as hospitalisation and death rate nor does it include the severity of different variants. If instead the aim of the testing scheme is to limit the death/hospitalisation rate then the testing policy in place may need to vary depending on risk. This means elderly people (and other groups at higher risk) would be more likely to be included in the pooled testing. This has been studied by other researchers for their individual testing policies but could be expanded to use pooled testing [Du et al. [2022]].

Pooled testing tends to work best in circumstances of low and relatively stable prevalence of the virus, as it means that a lot of the pools will come back negative. This reduces the number of individuals that have to be tested individually and so freeing up more tests for pools. However, low prevalence was not the case throughout the pandemic but in our current model, we are still using pooled testing. Therefore, our suggestion (work in progress) to overcome this problem is to have a ‘hybrid’ testing scheme where symptomatic individuals are still tested, but non-symptomatic individuals can be tested through pooled testing. By testing symptomatic individuals, it lowers the prevalence of the disease in the population that would be tested by pooled testing, hence making pooled testing more suitable. The different testing policies (i.e., whether both symptomatic and pooled testing would always be used or at certain points only symptomatic testing would be used and at others only pooled testing) and when to transition between them (i.e., for which performance metrics and for what values of these performance metric would the change occur) will be explored in a future paper.

## Data Availability

The basis of the code came from Johnson's code that is publicly available on his GitHub. Additional code will be made available on publication or beforehand upon request.

https://github.com/robj411/ADAGIO

## Acknowledgments

The authors would like to acknowledge Jack Bowden for helping to motivate this project. We would also like to acknowledge Anne Presanis and Shaun Seaman for their helpful comments on an earlier draft of this manuscript.

BH acknowledges the BSU for PhD funding.

SSV acknowledges funding and support from the UK Medical Research Council (MC UU 00002/15).

DSR received funding from the UK Medical Research Council (MC UU 00002/14).

This research was supported by the NIHR Cambridge Biomedical Research Centre (BRC1215-20014). The views expressed in this publication are those of the authors and not necessarily those of the NHS, the National Institute for Health Research or the Department of Health and Social Care (DHCS). For the purpose of open access, the author has applied a Creative Commons Attribution (CC BY) licence to any Author Accepted Manuscript version arising.

## Data Availability

The basis of the code came from Johnson [2023]’s code that is publicly available on his GitHub. Additional code will be made available on publication or beforehand upon request.

## A Appendix

The tables in the appendix contain some of the data that was used to make the graphs in the main text of the paper. All data used can be released upon request.

**Table 4:** Table of the expected value and standard deviation of total infections, length of epidemic, peak size, total isolations and the number of days the number of new infections surpasses five for symptomatic testing for varying non-compliance proportions for non-compliance type (a) for 200 replicates for 5 tests per day

| Non-complier Proportion (NCP) | Total Infections | Length | Peak Size | Total Isolations | Days over threshold |
| --- | --- | --- | --- | --- | --- |
| 0 | 369 (66.6) | 97.5 (13.8) | 56.1 (11.4) | 2410 (279) | 28.4 (8.18) |
| 0.05 | 371 (74.2) | 97.3 (13.5) | 57.0 (10.9) | 2350 (311) | 28.6(8.74) |
| 0.1 | 376 (67.4) | 101 (13.9) | 57.0 (10.2) | 2350 (275) | 28.7 (7.96) |
| 0.15 | 381 (64.7) | 103 (15.0) | 57.1 (10.6) | 2300 (259) | 29.1 (8.00) |
| 0.2 | 387 (66.7) | 107 (17.6) | 56.8 (11.0) | 2270 (271) | 29.6 (8.59) |
| 0.25 | 389 (70.0) | 106 (18.2) | 57.7 (11.7) | 2200 (267) | 29.6 (8.51) |

**Table 5:** Table of the expected value and standard deviation of total infections, length of epidemic, peak size, total isolations, total unneccesary isolations and the number of days the number of new infections surpasses five for pooled testing for varying non-compliance proportions for non-compliance type (a) for 200 replicates for 5 tests per day

| NCP | Total Infections | Length | Peak Size | Total Isolations | Unnecessary Isolations | Days over threshold |
| --- | --- | --- | --- | --- | --- | --- |
| 0 | 413 (42.4) | 116 (27.0) | 62.1 (10.6) | 6040 (873) | 4380 (635) | 31.6 (5.77) |
| 0.05 | 409 (40.3) | 116 (24.1) | 60.1 (10.3) | 5990 (855) | 4360 (597) | 31.2 (5.70) |
| 0.1 | 409 (40.1) | 118 (25.3) | 61.4 (10.3) | 6020 (856) | 4400 (609) | 31.0 (5.40) |
| 0.15 | 415 (41.3) | 118 (25.3) | 61.7 (9.85) | 6030 (808) | 4390 (601) | 31.9 (5.42) |
| 0.2 | 413 (41.0) | 113 (21.6) | 61.0 (9.16) | 5900 (756) | 4300 (558) | 31.9 (5.04) |
| 0.25 | 413 (41.8) | 116 (24.9) | 60.2 (10.1) | 5900 (851) | 4310 (620) | 31.9 (5.83) |

**Table 6:** Table of the expected value and standard deviation of total infections, length of epidemic, peak size, total isolations and the number of days the number of new infections surpasses five for individual random testing for for varying non-compliance proportions for non-compliance type (a) for 200 replicates for 5 tests per day.

| NCP | Total Infections | Length | Peak Size | Total Isolations | Days over threshold |
| --- | --- | --- | --- | --- | --- |
| 0 | 571 (50.3) | 143 (26.3) | 65.9 (10.4) | 161 (49.0) | 45.6 (6.18) |
| 0.05 | 568 (43.0) | 144 (25.8) | 66.1 (10.0) | 153 (43.2) | 45.1 (5.89) |
| 0.1 | 568 (44.9) | 142 (26.7) | 66.3 (10.6) | 158 (47.3) | 45.5 (6.00) |
| 0.15 | 569 (47.6) | 143 (24.9) | 66.6 (10.1) | 149 (50.9) | 45.7 (5.84) |
| 0.2 | 569 (42.5) | 143 (25.8) | 66.4 (10.4) | 156 (43.1) | 45.4 (5.47) |
| 0.25 | 570 (45.1) | 142 (24.1) | 66.2 (10.8) | 147 (47.7) | 45.6 (6.28) |
| Pooled random testing | 413 (44.6) | 112 (21.3) | 61.3 (9.59) | 6070 (832) | 32.0 (5.72) |
| individual random testing | 563 (45.9) | 143 (26.4) | 65.3 (10.3) | 153 (47.8) | 44.4 (5.83) |

**Table 7:**
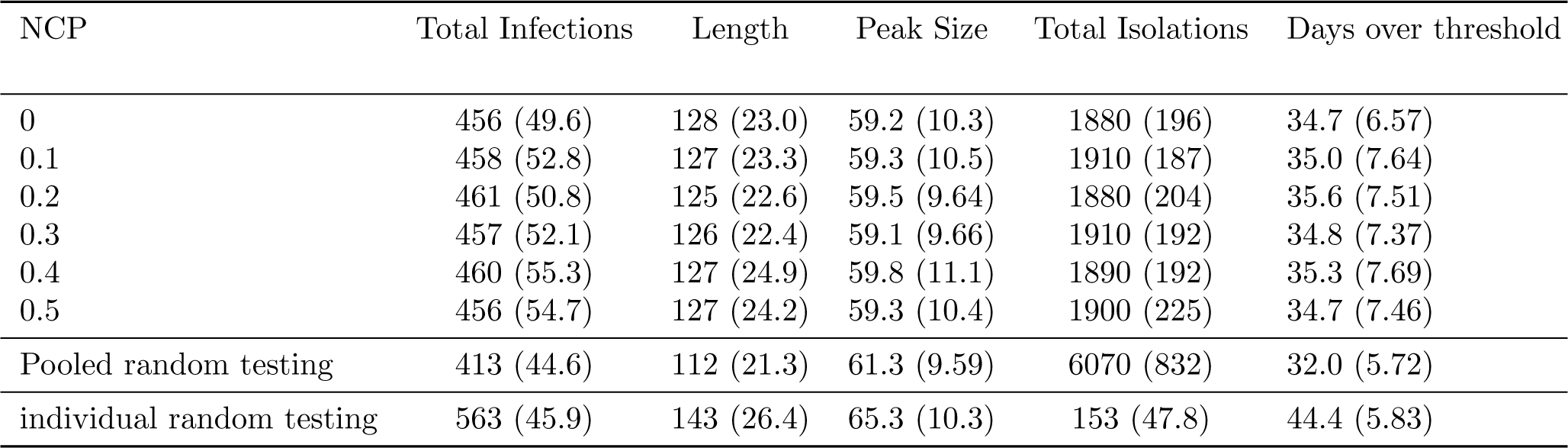
Table of the expected value and standard deviation of total infections, length of epidemic, peak size, total isolations and the number of days the number of new infections surpasses five for symptomatic individual testing for varying non-compliance proportions for non-compliance type (b) and with pooled testing and individual random testing as comparison for 200 replicates. This is for 5 tests per day and 0.02 proportion of non-complaince type (c).

## Notes

### Competing Interest Statement

The authors have declared no competing interest.

